# Syntax and Schizophrenia: A meta-analysis of comprehension and production

**DOI:** 10.1101/2024.10.26.24316171

**Authors:** Dalia Elleuch, Yinhan Chen, Qiang Luo, Lena Palaniyappan

**Author notes:** Address correspondence to: Dr. Lena Palaniyappan.

## Abstract

**Background:** People with schizophrenia exhibit notable difficulties in the use of everyday language. This directly impacts one’s ability to complete education and secure employment. An impairment in the ability to understand and generate the correct grammatical structures (syntax) has been suggested as a key contributor; but studies have been underpowered, often with conflicting findings. It is also unclear if syntactic deficits are restricted to a subgroup of patients, or generalized across the broad spectrum of patients irrespective of symptom profiles, age, sex, and illness severity.

**Methods:** We conducted a systematic review and meta-analysis, registered on OSF, adhering to PRISMA guidelines, searching multiple databases up to May 1, 2024. We extracted effect sizes (Cohen’s d) and variance differences (log coefficient of variation ratio) across 6 domains: 2 in comprehension (understanding complex syntax, detection of syntactic errors) and 4 in production (global complexity, phrasal/clausal complexity, utterance length, and integrity) in patient-control comparisons. Study quality/bias was assessed using a modified Newcastle–Ottawa Scale. Bayesian meta-analysis was used to estimate domain-specific effects and variance differences. We tested for potential moderators with sufficient data (age, sex, study quality, language spoken) using conventional meta-regression to estimate the sources of heterogeneity between studies.

**Findings:** Overall, 45 studies (n=2960 unique participants, 64·4% English, 79 case-control contrasts, weighted mean age(sd)=32·3(5·6)) were included. Of the patient samples, only 29·2% were women. Bayesian meta-analysis revealed extreme evidence for all syntactic domains to be affected in schizophrenia with a large-sized effect (model-averaged *d*=0·65 to 1·01, with overall random effects *d*=0·86, 95% CrI [0·67-1·03]). Syntactic comprehension was the most affected domain. There was notable heterogeneity between studies in global complexity (moderated by the age), production integrity (moderated by study quality), and production length. Robust BMA revealed weak evidence for publication bias. Patients had a small-to-medium-sized excess of inter-individual variability than healthy controls in understanding complex syntax, and in producing long utterances and complex phrases (overall random effects *lnCVR*=0·21, 95% CrI [0·07-0·36]), hinting at the possible presence of subgroups with diverging syntactic performance.

**Interpretation:** There is robust evidence for the presence of grammatical impairment in comprehension and production in schizophrenia. This knowledge will improve the measurement of communication disturbances in schizophrenia and aid in developing distinct interventions focussed on syntax - a rule-based feature that is potentially amenable to cognitive, educational, and linguistic interventions.

**Research in Context:** *Evidence before this study:* Prior studies have documented significant language deficits among individuals with psychosis across multiple levels. However, syntactic divergence—those affecting sentence structure and grammar—have not been consistently quantified or systematically reviewed. An initial review of the literature indicated that the specific nature and severity of syntactic divergence, as well as their impact on narrative speech production, symptom burden, and daily functioning, remain poorly defined. We conducted a comprehensive search of the literature up to May 1, 2024, using databases such as PubMed, PsycINFO, Scopus, Google Scholar, and Web of Science. Our search terms combined psychosis, schizophrenia, language production, comprehension, syntax, and grammar, and we identified a scarcity of meta-analytic studies focusing specifically on syntactic comprehension and production divergence in psychosis.

*Added value of this study:* This systematic review and meta-analysis is the first to quantitatively assess syntactic comprehension and production divergence in individuals with psychosis. This study provides estimated effect sizes associated with syntactic impairments as well as a quantification of the variance within patient groups for each domain of impairment. Besides a detailed examination of this under-researched domain, we also identify critical research gaps that need to be addressed to derive benefits for patients from knowledge generated in this domain.

*Implications of all the available evidence:* This study provides robust evidence of grammatical impairments in individuals with schizophrenia, particularly in syntactic comprehension and production. These findings can enhance early detection approaches via speech/text readouts and lead to the development of targeted cognitive, educational, and linguistic interventions. By highlighting the variability in linguistic deficits, the study offers valuable insights for future therapeutic trials. It also supports the creation of personalized formats of information and educational plans aimed at improving the effectiveness of any therapeutic intervention offered to patients with schizophrenia via verbal medium.

## 1. Introduction

The cognitive faculty of language supports interpersonal communication and thinking^1^, both of which are disrupted in psychotic disorders such as schizophrenia. Do the thought and communication disorders in people with schizophrenia result from structural issues i.e., grammatical impairment (syntactic divergence) in people with schizophrenia? This question has been studied at various times in the past, with a variety of methods and approaches ^2–6^. Despite the substantial body of work, the existing literature presents a fragmented understanding of the nature and extent of syntactic deficits. Disorganised speech, a diagnostic feature of schizophrenia in DSM-5 ^7^, is assessed on the basis of incoherence that leads to a failure of effective communication. Syntax production, if impaired, can generate conversational incoherence. Similarly, impaired comprehension of syntax (i.e., who did what to whom?) may contribute to impaired meaning and misinterpretations that typify positive psychotic symptoms such as persecutory delusions. In the current study, we systematically review the literature published to date on both syntactic production and comprehension in schizophrenia.

Producing and inferring meaning via language is not based on isolated lexical concepts (semantic categories), but involves the interactional basis offered by grammatical constructions. Grammar enables the signifiers and the signified to be put together. Thus, there is a strong case to be made for syntax-level deficits i.e., an aberration in the way words are put together, to have primacy in the language disorder of schizophrenia ^8–11^. Several thoughtful reviews in recent times have hinted at the critical importance of syntactic divergence in schizophrenia ^4,12–15^. Bora and colleagues highlighted a role for syntactic comprehension divergence when analyzing the linguistic correlates of the burden of formal thought disorder^16^. Nonetheless, to our knowledge, a comprehensive meta-analytic quantification of the overall magnitude of grammatical impairment in both comprehension and production in schizophrenia is still lacking.

Quantifying the degree of grammatical impairment in schizophrenia is critical for two reasons. Firstly, the use of the various linguistic markers in speech to predict clinically important outcomes is an emerging pursuit in the field (e.g., onset of first episode ^17–19^, relapses ^20^). Despite the many studies carried out to date, one major obstacle in bringing such predictive analytics to routine clinical use is the lack of empirical guidance on feature selection in these models. As a result, a large number of automatically derived linguistic variables are being tested in clinical prediction models, with minimal overlap among different studies, impeding interpretability and successful external validation (e.g., not a single linguistic feature overlapped across the 18 prediction analysis studies identified in a recent review^21^). This can be addressed via evidence-based preselection of variables that most proximally relate to the clinical construct of interest i.e., the presence of schizophrenia in our case [see Meehan and colleagues^22^ for a state-of-the-art review]. Meta-analytic estimation of the effect size of syntax production/comprehension variables will provide evidence for their utility in speech-based predictive analytics.

Secondly, given the relevance of social interaction for functional recovery^23^, interventions to ameliorate communication deficits in schizophrenia are steadily growing in recent times ^24–26^. Outcomes of these trials can be improved by identifying the most affected syntactic markers as treatment targets and identifying if distinct subgroups with varying degrees of deficits are likely to occur among patients. In the presence of a high degree of interindividual variability in syntactic deficits, stratified RCTs for communicative remediation are likely to have a better yield. Thus, meta-analytic estimation of the effect size and variability of syntactic deficits will inform forthcoming intervention trials.

Our primary goal of this review is to provide a quantitative synthesis of the degree and interindividual variability of syntactic language deficit across the domains of syntactic comprehension, anomaly/error detection, and various levels of complexity and integrity of syntactic production in schizophrenia. We also aim to investigate the relationship between syntactic production, comprehension, and symptom severity and identify potential research gaps and opportunities in this area of work.

## 2. Methods

### 2.1 Search Strategy and Selection Criteria

The original protocol was registered on the Open Science Framework registry (May 202), with an update after the initial search but before undertaking statistical analysis (October 2024)^27^. This update included missing information on meta-analytic methods and bias assessment framework, adding specifications (grouping of syntactic domains, metaregression variables) and planned deviations (reporting pronoun aberrations separately from the current report, dropping reaction time and parts-of-speech measures to reduce bias from reporting inconsistencies). Any further deviations that occurred after the data-analysis (the use of multivariate approach to meta-analysis) are reported as Supplemental Results. This review adheres to the Preferred Reporting Items for Systematic Reviews and Meta-Analyses (PRISMA) guidelines^28^ and recent recommendations to protect against researcher bias in meta-analysis^29^. We performed a literature search across multiple electronic databases, including PubMed, PsycINFO, Scopus, and Web of Science, up to May 1, 2024. Search terms included a combination of keywords and Medical Subject Headings (MeSH) terms related to schizophrenia (schizophrenia OR schizo* OR psychos* OR psychot*), language (language OR verbal OR linguistic OR speech OR communicat* OR thought), syntax (syntax OR syntactic OR gramma*). Two reviewers (DE and LP) independently screened titles and abstracts against the inclusion criteria using Rayyan software after removing duplicates. Full texts of relevant studies were assessed for eligibility. We then added further studies to the pool by screening the bibliography and hand-searching all citations received by the identified studies via Google Scholar.

We included English/French language publications describing studies that (1) enrolled adults (aged 18 or above) diagnosed with schizophrenia spectrum disorders (schizophrenia, schizoaffective, or schizophreniform psychosis) and a control group of healthy adults without known psychiatric disorders (2) assessed speech production and/or comprehension, focusing on grammar and syntax. This includes evaluating either grammatical comprehension (by quantifying a person’s ability to *understand complex sentences* or *detect errors* in the syntactic formation) and/or production (by assessing the degree of global [narrative level] or local [clausal/phrasal level] complexity, length and integrity in the utterances or sentences). This grouping of domains of interest was based on Morice and Ingram’s original work^30^ that separated *complexity* and *integrity* in syntax production in schizophrenia, with *phrasal/clausal level complexity* (coordination) later included by Thomas and colleagues^31^. This set was further extended as per Lu’s Syntactic Complexity Analyzer approach^32^ to distinguish *production length* from other complexity measures.

Only empirical studies with quantitative measures derived in the same manner from both groups were included. Studies focused on subjects <18 years of age^33–35^, case reports/case series^36^, and those without a healthy control group^11,37–41^ were excluded. Additionally, studies focussed on high-risk subjects without a diagnosed schizophrenia spectrum disorder^42^, studies reporting verbal outputs that were either restricted (e.g., scripted conversations^43^) or likely to have been edited after production (e.g., written reports and social media texts^44–48^), non-naturalistic speech (e.g., word list generation, repetition, monitoring or recall of memorized text^49–51^), analysis restricted to parts-of-speech tagging (with no sentential syntax)^52–55^ or providing only second order derivatives (e.g., speech graph metrics^56^ or factor scores^57^) without direct indices of syntax production/comprehension were not eligible. One study with a retraction notice was also excluded. Studies with unconventional criteria for syntactic complexity^58–60^ and those without quantitative measures or plots that allowed effect size estimation were also excluded^49,61,62^. For a list of articles excluded at the stage of data extraction with the reasons for exclusion and main results, see Supplemental Table 1.

### 2.2 Data Extraction

We extracted the available clinical/demographic data (author(s), publication year, country, sample size, mean age, and symptom severity based on standardized scales [e.g., PANSS, SANS/SAPS, BPRS/BPRS-E, with the reported total scores in each patient sample converted to a scale of 0 to 1 via min-max transformation (See Supplement)], sex distribution, chlorpromazine equivalent of antipsychotic dose (conversions from other drug equivalents or Defined Daily Doses as per^63^), mode [free speech, visual/verbal stimulus such as picture/proverb elaboration, sentence to picture matching] and the language of task administration). A description of the included studies is available in Table 1. When overlapping samples were published in more than one paper, we extracted data from the largest reported sample^64,65; 66,67;68,69;30,70;71,72^. We reached out to selected authors (12·2%) when quantitative measures were unclear for clarifications. For studies where numerical values were not provided^31,57,73–76^ we extracted these values from published plots using a visual data extraction tool (plotdigitizer.com). When more than one mean was reported on the same measurement from the same sample (e.g., on/off medications as in^77^), we included the average as the summary measure. Some of the studies reported median and range values instead of mean and SD required for Cohen’s *d* estimation^51,74,78^. In such instances, we used the five-number summary approach^79^, available at https://www.math.hkbu.edu.hk/~tongt/papers/median2mean.html.

**Table 1.**
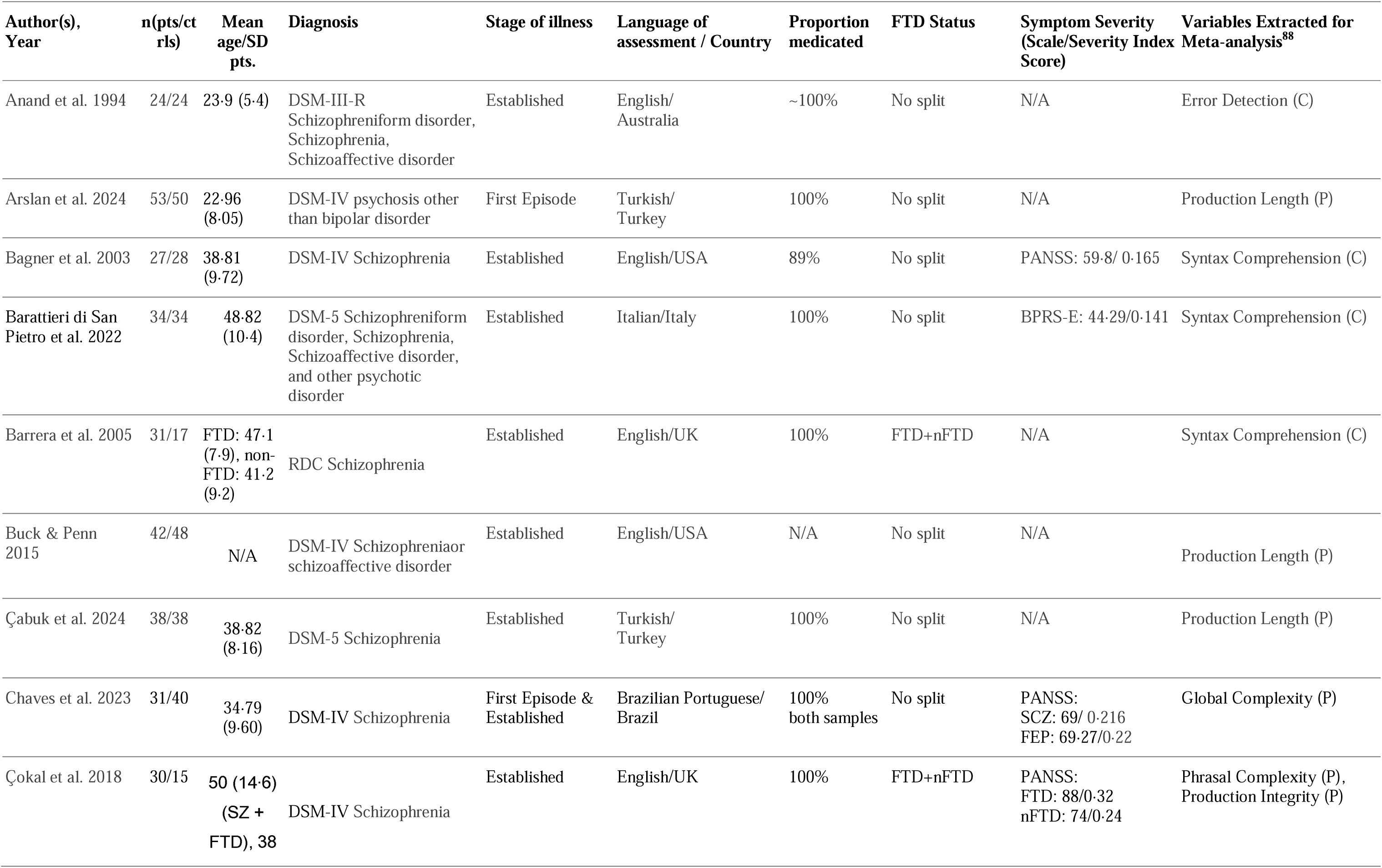
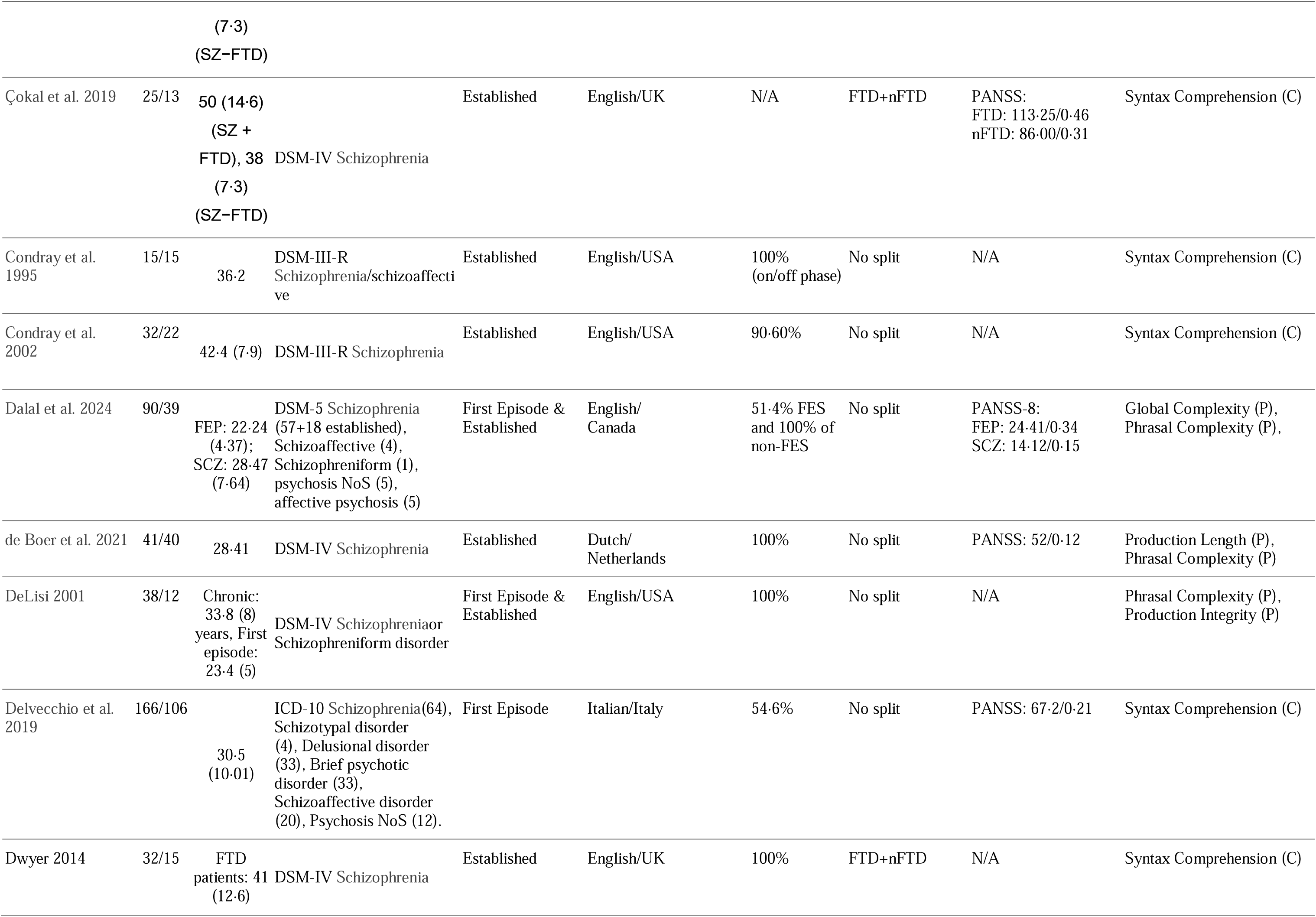
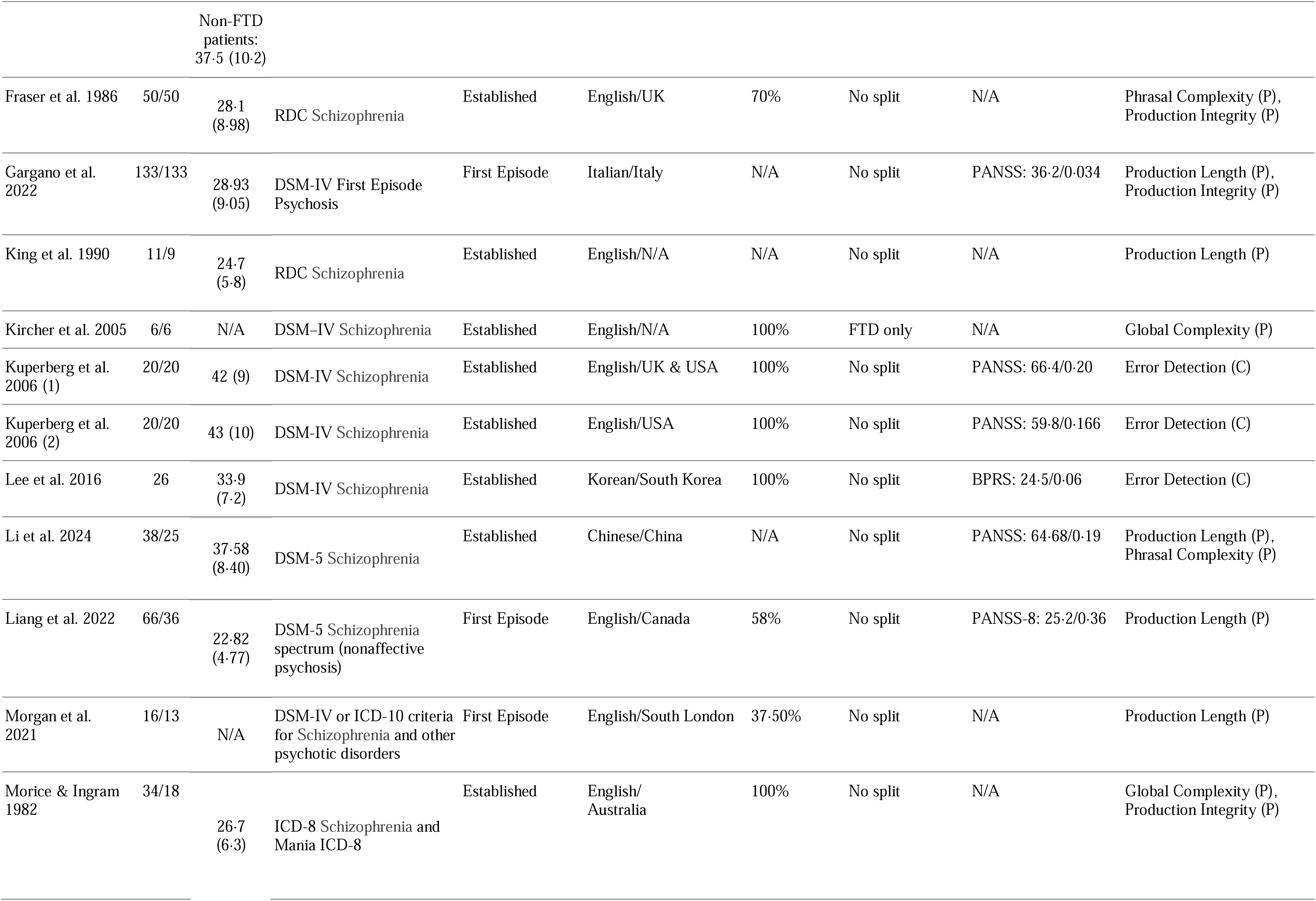
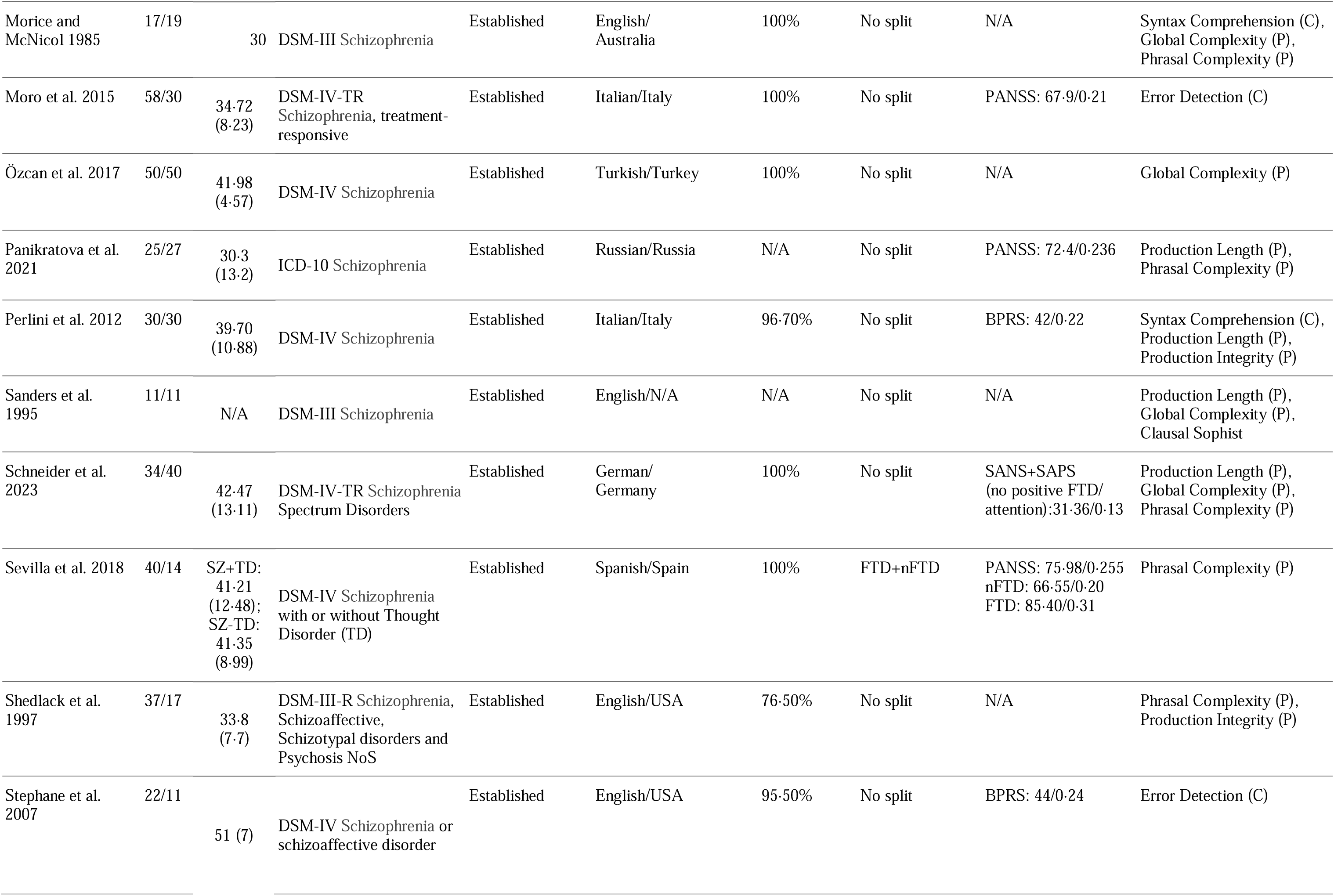
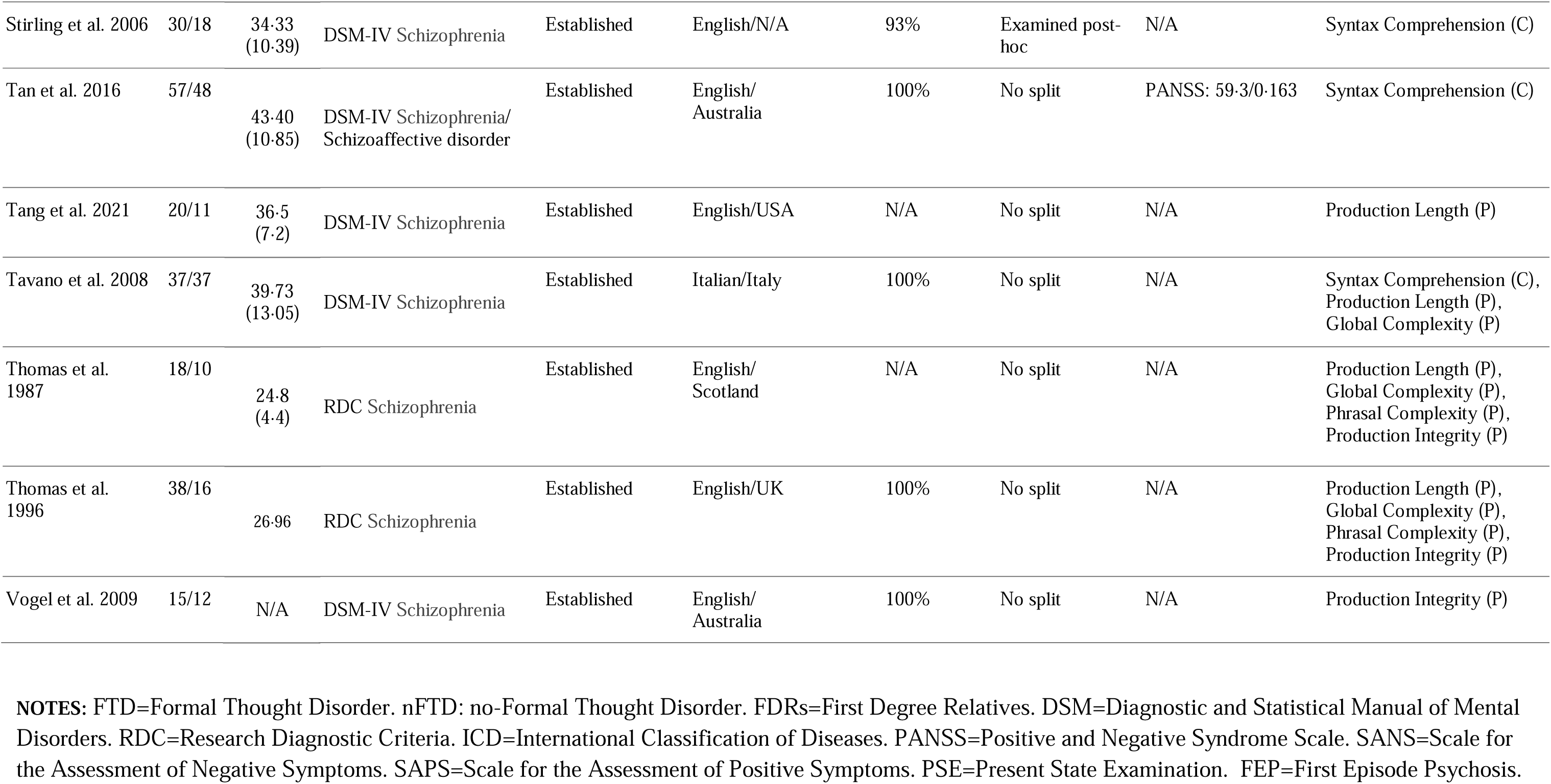
Description of the included studies.

### 2.3 Quality Assessment

The quality of the studies was assessed using a purposively modified Newcastle–Ottawa Scale^80^, widely used in psychiatry where rating scale use for exposure assignment is a common practice^81^. The following indicators were evaluated: case definition, representativeness, selection of control group, comparability of groups, ascertainment of ‘exposure’ (i.e., measurement of syntactic variables of interest), and quality of data reporting. Items in the Newcastle-Ottawa framework are known to have low reliability among raters^82^ (e.g., demonstrating the timing of measurements) and lack of clarity^83^ (e.g., emphasis on independent validation of the case status, response proportions, the practice of higher scores for population-based controls, statistical adjustment and blinding which are often unsatisfactory in case-control designs) were replaced these with items specific to psychiatric diagnoses and linguistic variable assessment (see Supplemental Table 2 for a description). Furthermore, we defined likely confounders a priori for bias assessment (age, sex, education, and native language being different from the language of assessment). Each study was independently rated for scores between 0-12 by two authors (DE and LP), with disagreements resolved by discussion.

### 2.4 Data Analysis

Statistical analyses were conducted using the JASP 0.19.0.0 package^84^. Effect sizes were calculated from available means and standard deviations (Cohen’s d = (M_2_ - M_1_) ⁄ SD_pooled_) from each set of analysis. As some studies reported error rates while others reported accuracy rates, all effect sizes were sign-adjusted to read as controls>patients when producing summary values.

We pooled the *d* values using bayesian model-averaged (BMA) meta-analysis via metaBMA R package implemented in JASP^85^. BMA evaluates the likelihood of the data under a combination of models regarding the meta-analytic effect and heterogeneity, reporting model-averaged effects. Evidence in favor of a group difference was categorized as weak (for BF_10_ 1 to <3), moderate (BF_10_ 3 to <10), strong (BF_10_ 10 to <30), very strong (BF_10_ 30 to <100), and extreme (BF_10_ >100).

Meta-regression analyses were performed when sufficient evidence for heterogeneity between studies was uncovered in any domain. We included the mean age of patients, proportion of female patients, mean chlorpromazine equivalent dose, language of the study assessment (English vs. non-English), and study quality scores as potential moderators.

Robust Bayesian meta-analysis^86^ was used to assess the sensitivity of the results to the potential presence of publication bias and heterogeneity.

Log Coefficient of Variation Ratio^87^ (lnCVR: natural log of ratio of the estimated total coefficient of variation between the patient and the control group) was used to quantify the difference in variability after scaling to the mean of each group [lnCVR= 0 indicates equal variability; >0 greater variability, while <0 indicates lower variability in patients vs. controls].

Given the between-domain heterogeneity, we used a random-effects model to pool the 6 *lnCVR* measures and the 6 Cohen’s *d* estimates across the domains to assess the overall effect.

## 3. Results

### 3.1 Study Selection

A total of 463 studies were identified through the initial database search. After removing duplicates, 289 unique studies remained. Following title and abstract screening, 86 articles were retrieved as relevant, of which 45 studies met the inclusion criteria for numerical synthesis for the meta-analysis^88–95,77,96,97,64,9,67,98,51,99–104,73,78,105,74,30,106–114,75,115–118,71,31^ (see Figure 1).

**Figure 1:**
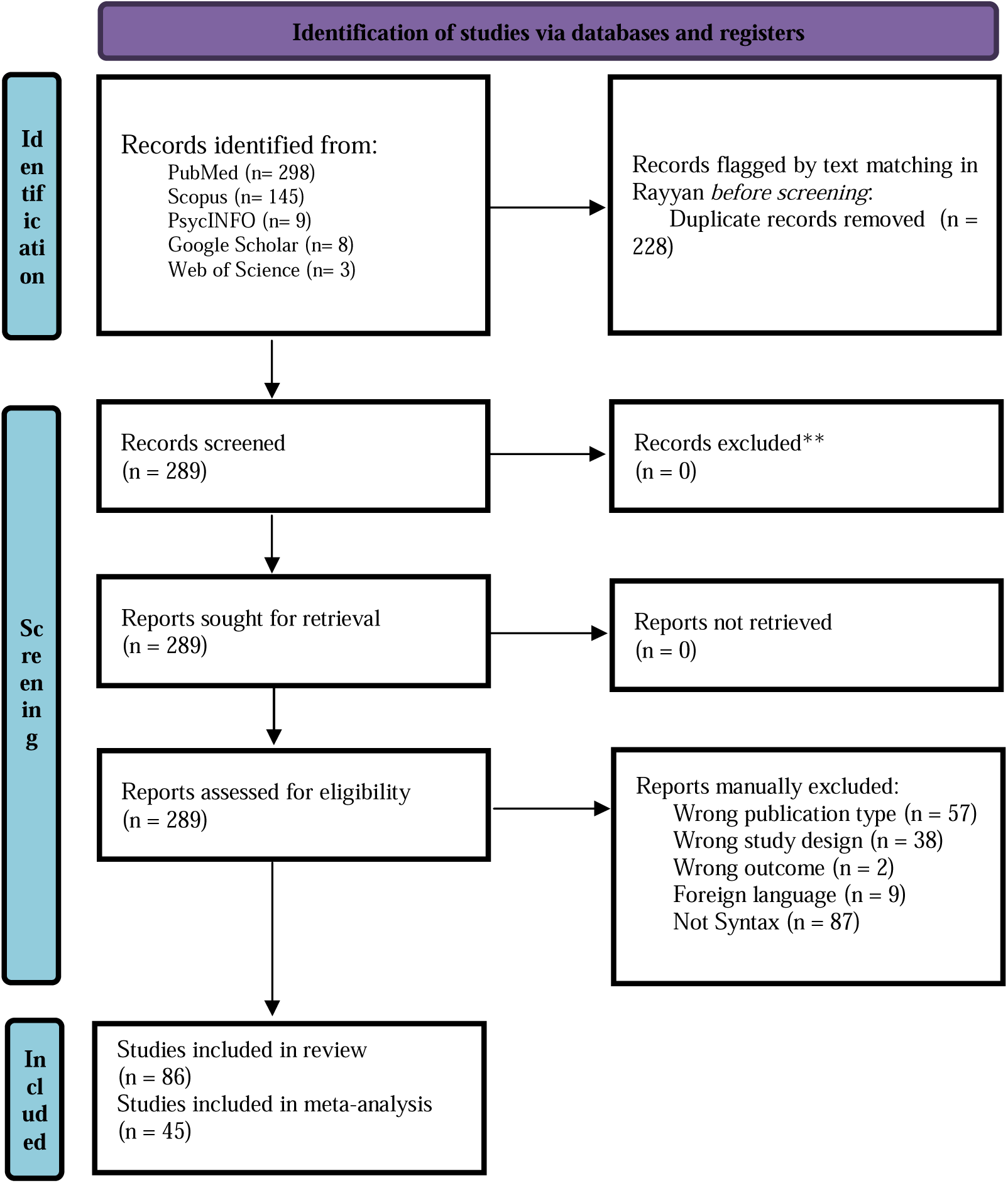
**PRISMA 2020 flow diagram for the systematic review of syntax and schizophrenia.**

### 3.2 Study Characteristics

The final list included studies published between 1982 and 2024, with summary data from a total of n= 1679 patients and n= 1281 controls available from 79 comparisons across 6 domains of interest. The weighted mean age across studies was 32·31 (SD=5·6) years, with no difference in distribution among patients and control cohorts (paired t=0·85, p=0·4). Only 29·2% of participants were women, with 5 studies recruiting only men^77,102,109,119,120^; only 8 studies had >40% women. The studies predominantly included individuals diagnosed with established schizophrenia spectrum disorders (n=1292), with first-episode samples forming 33·59% of the total sample (n=564). A great majority of studies (64·4%) recruited English-speaking participants. Some studies reported separate contrasts based on the presence of Formal Thought Disorder (FTD/no-FTD^121–123^) or stage of illness (FEP/established schizophrenia^9,64,95^). Quality scores are presented in Supplemental Table 3.

There is no single accepted index to measure grammatical impairment in mental health conditions. As a result, we found a notable variation in the method used to quantify the variables of interest, and in some cases, more than one variable for the same domain was reported. As a general principle, we chose the measures with the closest theoretical alignment to the 6 domains of interest for this meta-analysis. Within each domain, we chose tasks and variables that were most commonly used across studies. Other study-specific decisions in variable choices are discussed in the Supplement.

### 3.3 Data availability

While mean age (93·3% of studies), language of testing (100%), and sex distribution (93·3%) were available for most studies, an estimate of antipsychotic dose exposure (48.9%) and overall symptom severity (40%) were less often reported. Most studies only provided the overall proportion of antipsychotic use and domain-specific symptom scores (generally positive symptoms). As a result, we included age, assessment language, sex, and the study quality scores in the meta-regression analyses, but only reported moderator/effect-size bivariate correlations for antipsychotic dose and total symptom severity index.

### 3.4 Meta-analytical results

The results of Bayesian Meta-Analysis for each group of studies are shown in Table 2 along with the data on between-studies heterogeneity, log coefficient of variation ratios, and publication bias. BMA showed extreme evidence for reduced syntactic comprehension, error detection, production length, phrasal complexity, production integrity, and global complexity in patients (all BF_10_>100; Figure 2). Random effects analysis across the 6 domain-specific effects indicated extreme evidence (BF_10_=3173; estimated *d*=0·87) for an overall grammatical impairment in schizophrenia. See the supplement for multivariate meta-analysis of correlated outcomes.

**Figure 2:**
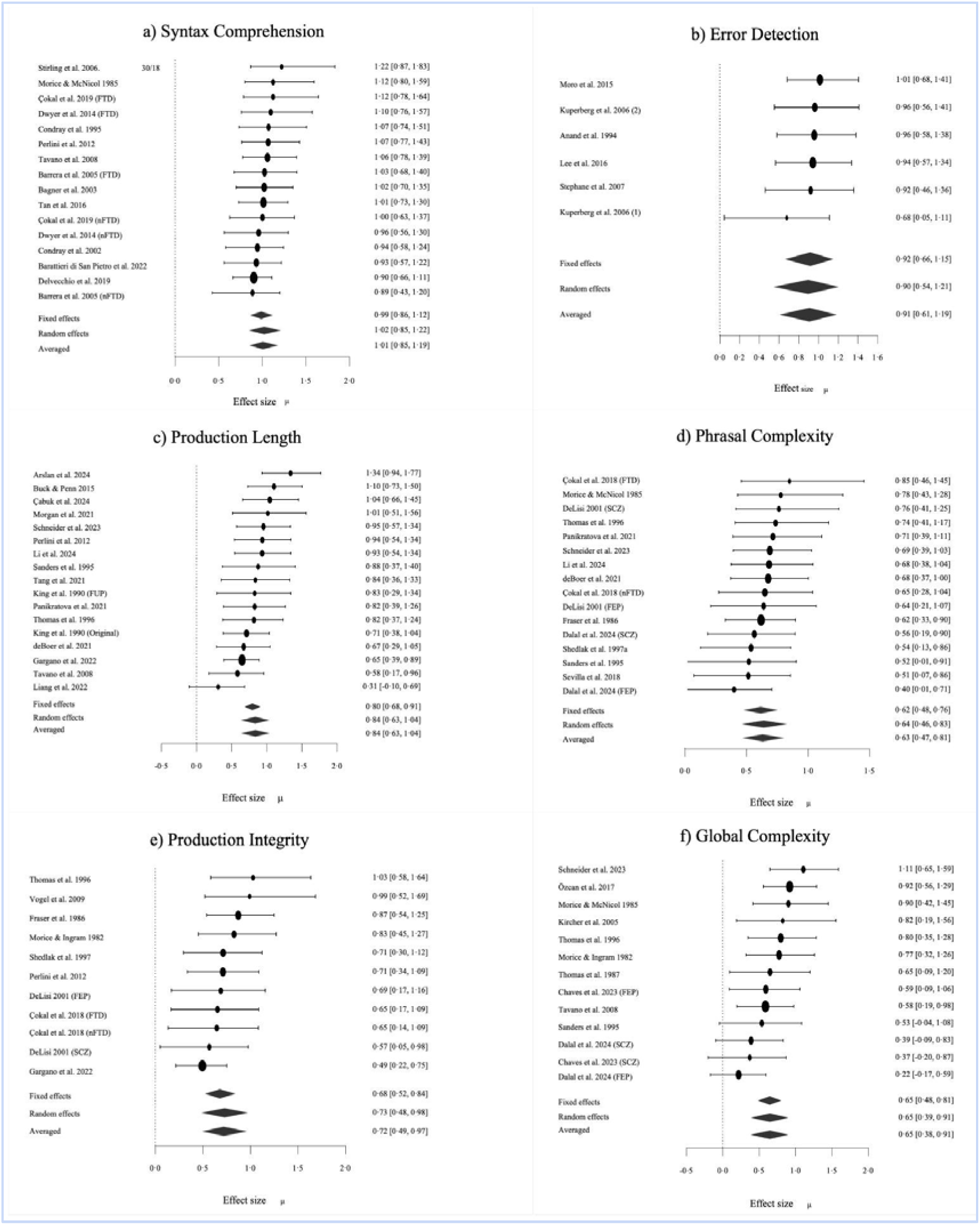
Forest plots for domain specific meta-analyses of syntactic production and comprehension in schizophrenia. Bayesian Model Averaged estimates of group differences in syntactic comprehension (k=16), error detection (k=6), production length (k=17), phrasal complexity (k=16), production integrity (k=11) and global complexity (k=13). Estimated (not observed) Cohen’s d values are given along with 95% credible intervals in parentheses. FTD=Formal Thought Disorder. nFTD: no-Formal Thought Disorder. FEP=First Episode Psychosis. SCZ=Established schizophrenia

**Table 2:** Summary of Bayesian Model-Averaged Meta-Analysis of Grammatical Impairment in Schizophrenia Spectrum Disorders.

| Syntactic Domain | k | N Patients, Controls | Cohen's d (BMA:95% CrI) | BF<sub>10</sub> for H1 | Heterogeneity Tau (95% CrI) | BF<sub>rf</sub> for RE | Significant Moderator effect (s.e.) | lnCVR (BMA:95% CrI) | ROBMA BF<sub>10</sub> for publication bias & mean difference |
| --- | --- | --- | --- | --- | --- | --- | --- | --- | --- |
| Syntactic Comprehension | 16 | 530/400 | 1.01 [0.85,1.19] * | 220x10<sup>5</sup> Extreme | 0.18 [0.04,0.42] | 1.08 Weak | None | 0.41 [0.11,0.71]* | Weak publ. bias (1.53)<br>Extreme effect (244) |
| Error Detection | 6 | 170/134 | 0.91 [0.60,1.19] * | 303 Extreme | 0.21 [0.04,0.61] | 1.01 Weak | None | 0.25 [-0.52,0.96] | No publ. bias (0.72)<br>Strong effect (23.0) |
| Production Length | 17 | 646/614 | 0.84 [0.63,1.04] * | 460 x10<sup>3</sup> Extreme | 0.31 [0.15,0.53]* | 313.75 Extreme | None | 0.13 [0.00,0.25]* | Weak publ. bias (2.78)<br>Strong effect (7.78) |
| Phrasal Complexity | 16 | 489/325 | 0.63 [0.46,0.81] * | 335 x10<sup>2</sup> Extreme | 0.20 [0.05,0.45] | 1.78 Weak | None | 0.29 [0.03,0.56]* | Weak publ. bias (1.41)<br>Moderate effect (3.52) |
| Production Integrity | 11 | 368/303 | 0.73 [0.49,0.99] * | 1258 Extreme | 0.27 [0.06,0.60]* | 4.67 Strong | Quality* 0.31(0.12) | 0.12 [-0.08,0.32] | No publ. bias (0.96)<br>Strong effect (9.95) |
| Global Complexity | 13 | 335/246 | 0.65 [0.39,0.92] * | 362 Extreme | 0.35 [0.14,0.65]* | 54.86 Very Strong | Age<sup>a</sup>* 0.04(0.01) | 0.22 [-0.01,0.43] | Weak publ. bias (1.06)<br>Moderate effect (4.42) |

a. Estimated from k=12; higher deficits in samples with higher mean age. \* p<0.05. ROBMA: Robust Bayesian meta-analysis s.e. standard error K=number of studies. N=sample size based on unique participant counts CrI=Credible Intervals. BF<sub>rf</sub> = Bayes Factor for random effects over fixed effects. BF<sub>10</sub> = Bayes Factor for evidence for the presence of expected group differences over null hypothesis of no difference. BMA: Bayesian Model Average. lnCVR = natural log of coefficient of variation ratio for patients vs. controls RE: random effects. Coloured boxes indicate significant results as per frequentist statistics (p<0.05).

**Table 3.**
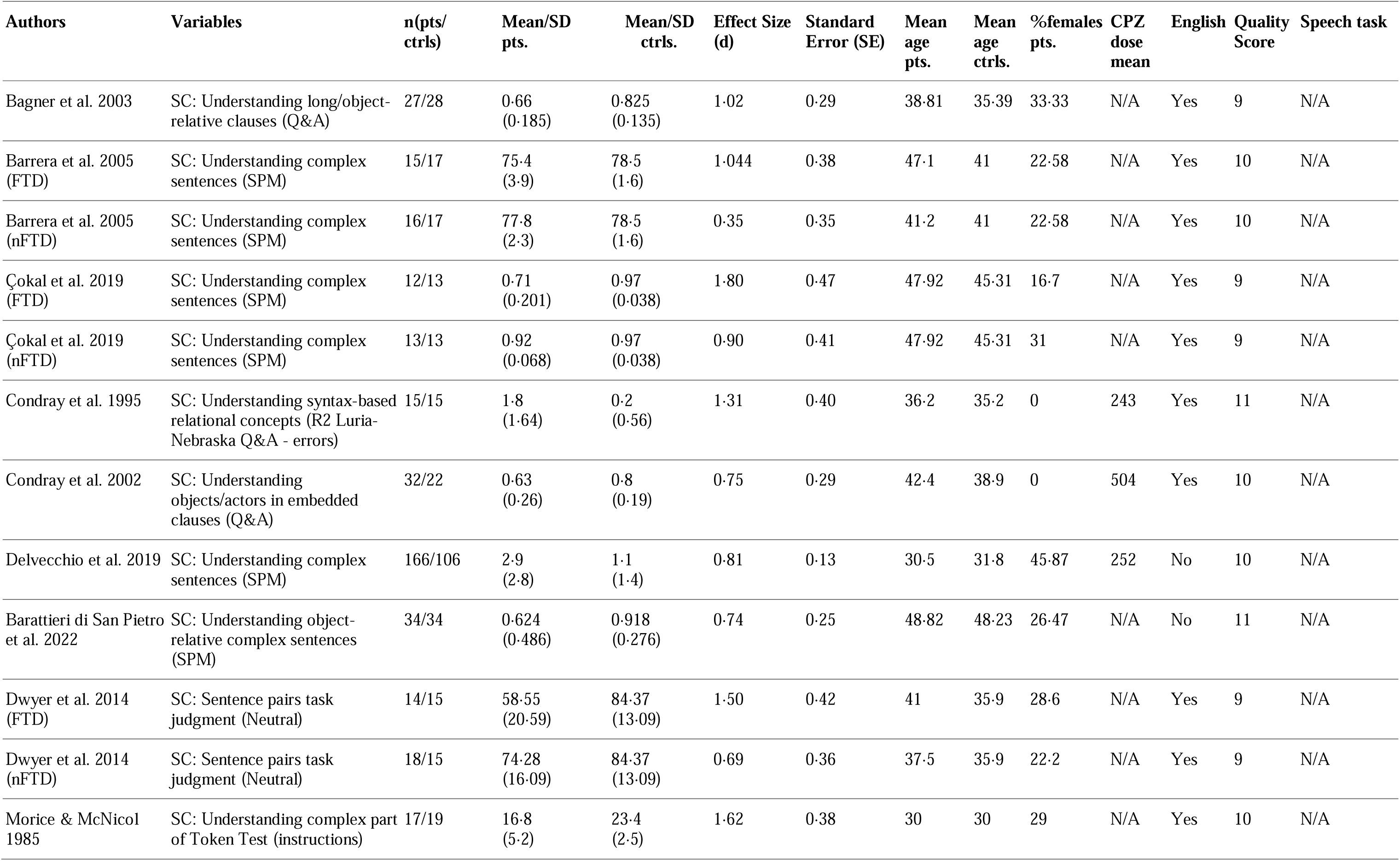
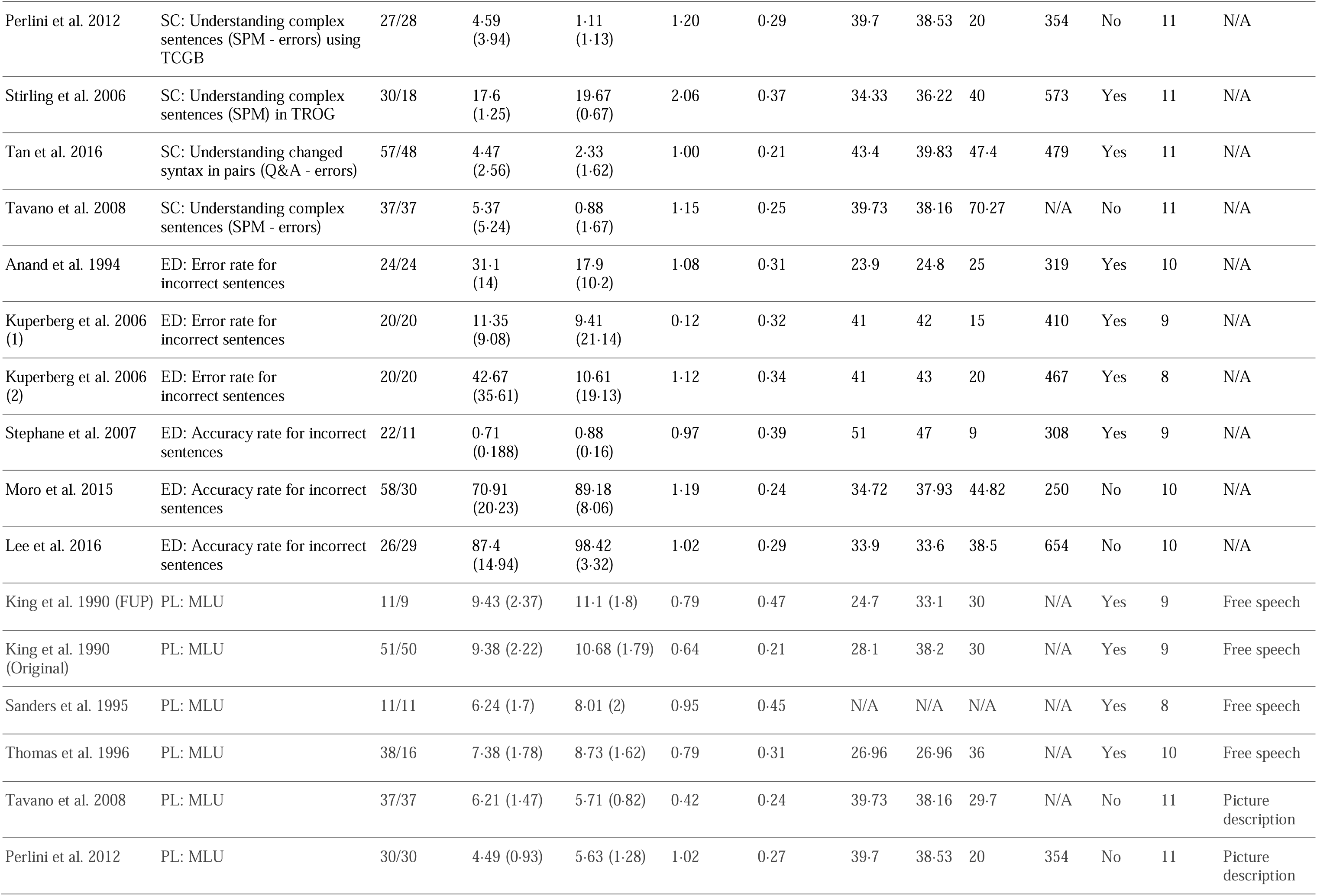
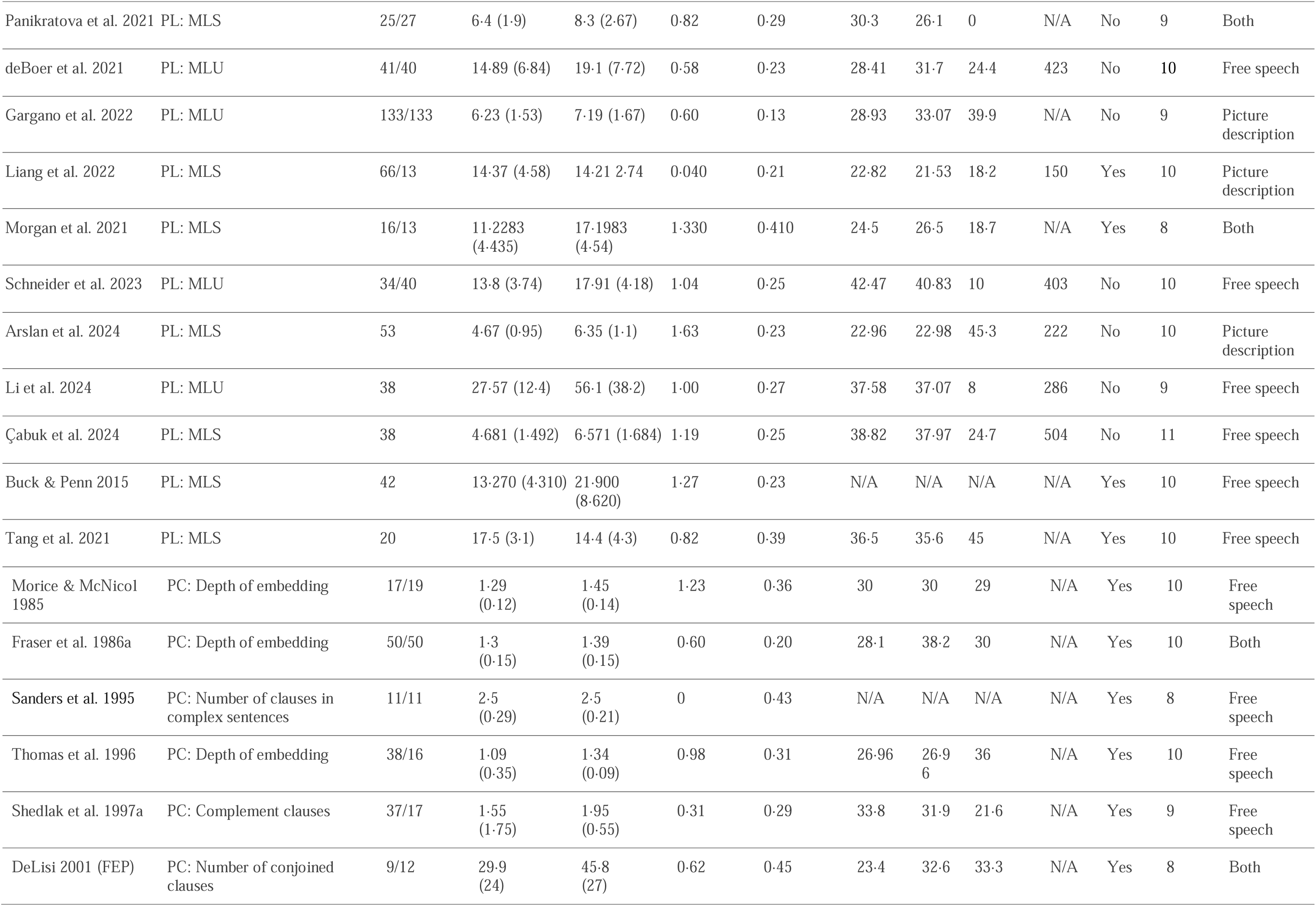
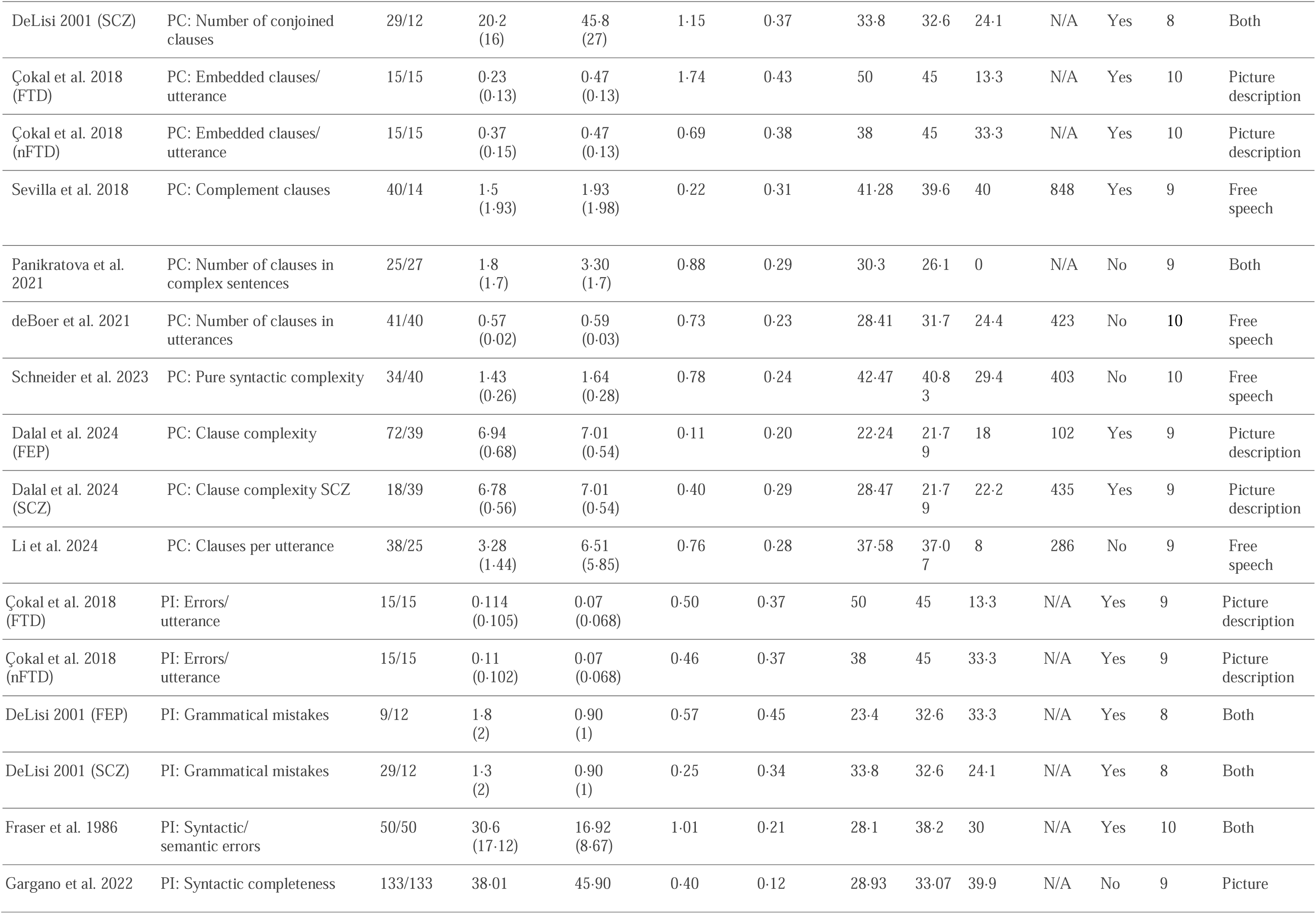
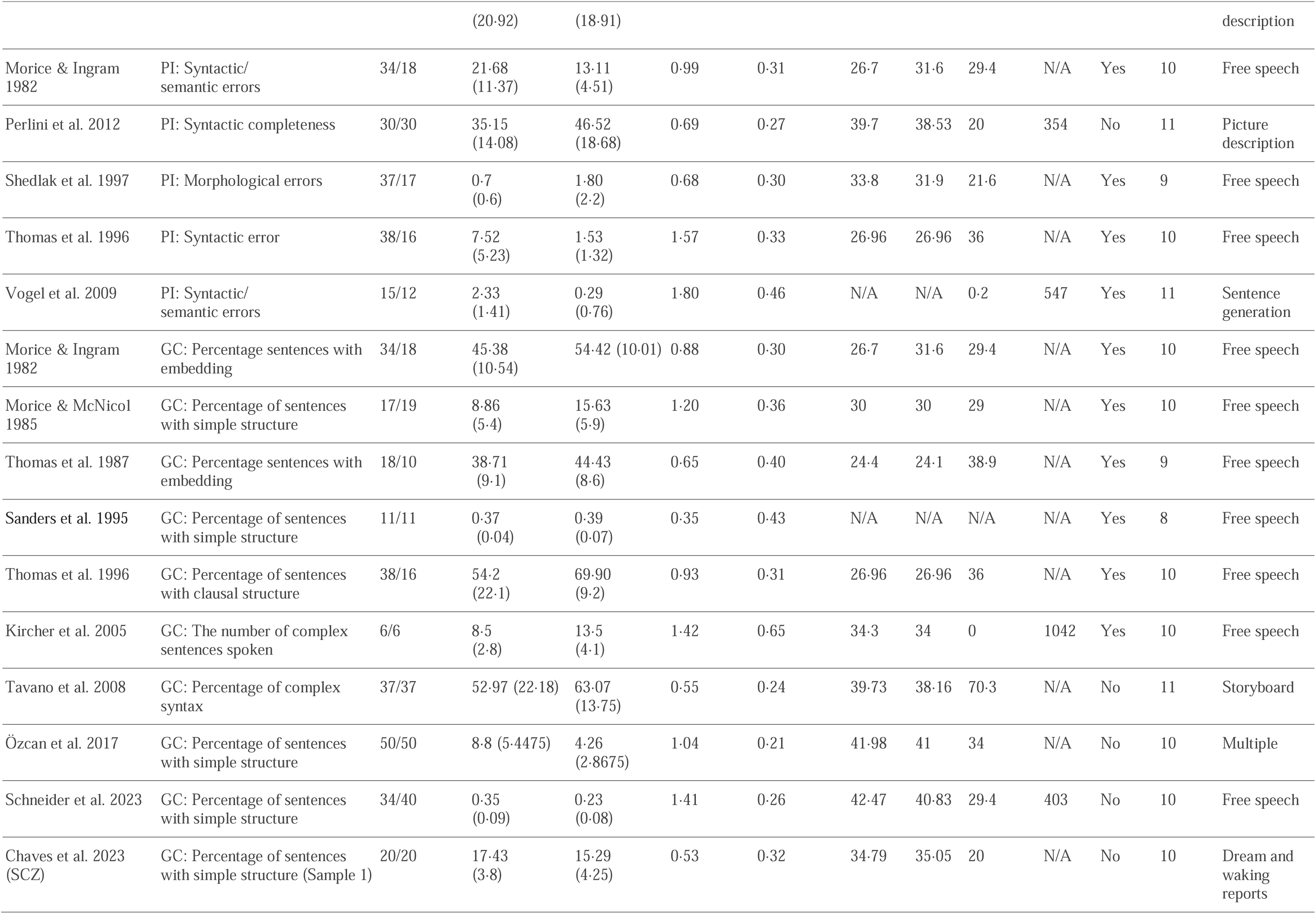
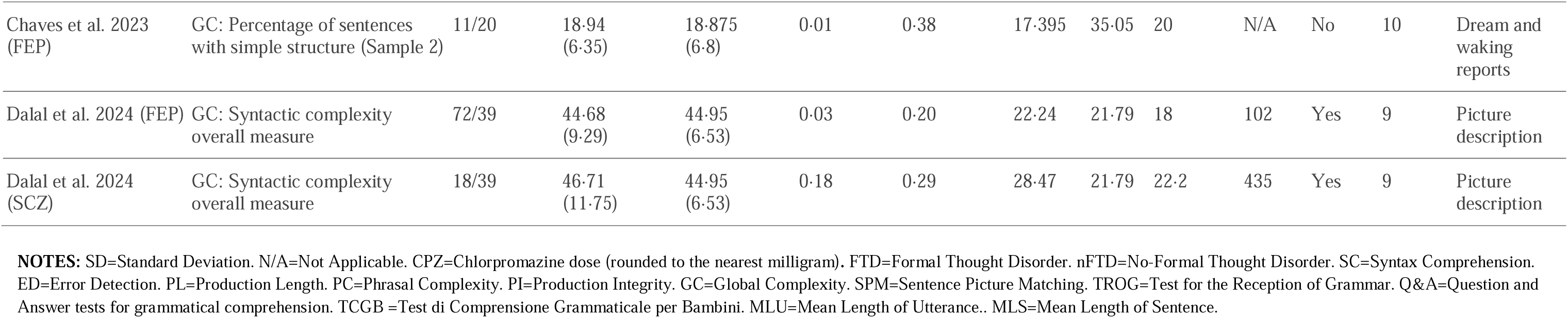
Key variables and moderators for the case-control comparisons in the meta-analysis.

Between study heterogeneity (tau) was strong for global complexity, production length, and integrity. Of these domains, the meta-regression analysis revealed age as a significant moderator for global complexity while study quality was the most significant known source of heterogeneity for production integrity (Table 2; Fig 3). The moderator/effect-size bivariate correlations were not significant for antipsychotic dose (r_31_=0·27, p=0·14) or total symptom severity index (r_33_=0·06, p=0·74) across all domains. While the number of studies on clinically detectable FTD was insufficient for a meta-regression, visual inspection of the forest plots revealed that all FTD contrasts had above-average Cohen’s *d* values for syntactic comprehension and phrasal complexity but not for production integrity.

**Figure 3:**
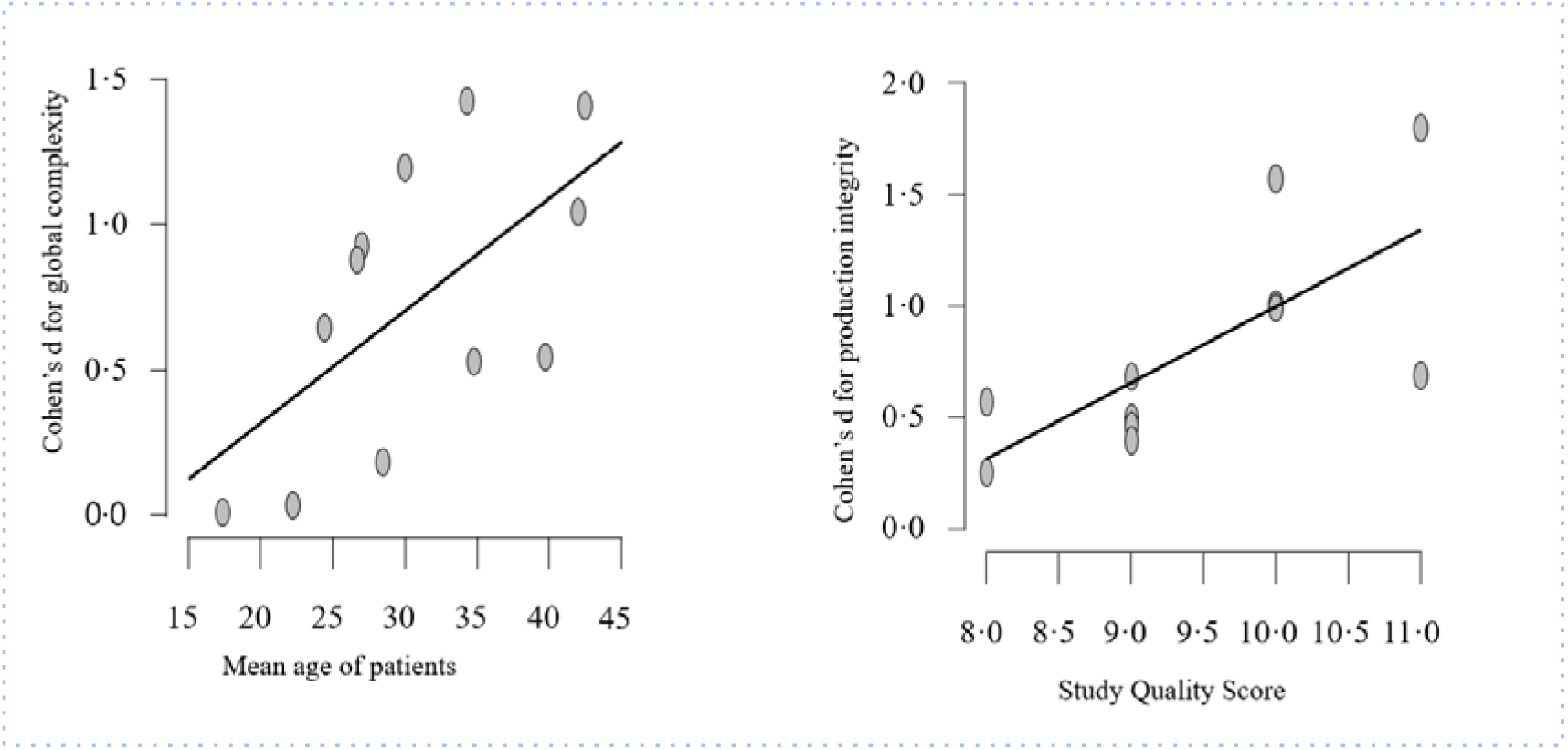
Observation from meta-regression analysis. Older study cohorts had more pronounced effect size differences for global syntactic complexity while better quality studies reported higher effect sizes for production integrity

Meta-analysis of within-group variations indicated higher inter-individual variability in patients for syntactic comprehension, phrasal complexity, and production length (lnCVR= 0·13-0·41; medium to large variation effect^124^) but not for other measures (Figure 3). Random effects analysis across the 6 domain-specific variation estimates indicated moderate evidence (BF_10_ = 5·27; estimated lnCVR = 0·21) for excess variability among patients compared to healthy controls (Figure 4).

**Figure 4:**
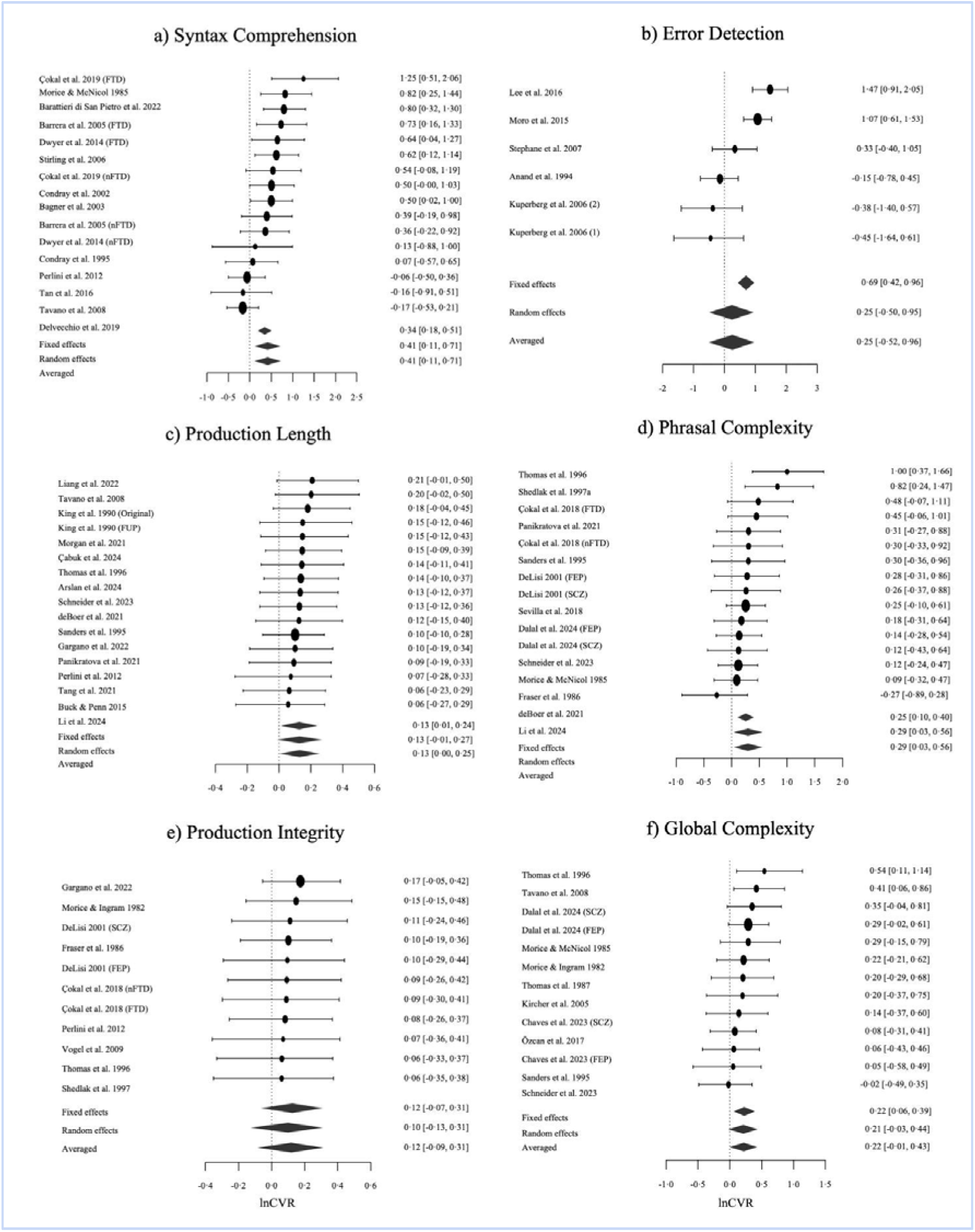
Forest plots for domain specific meta-analyses of variation in syntactic production and comprehension in schizophrenia. Bayesian Model Averaged estimates of group differences in syntactic comprehension (k=16), error detection (k=6), production length (k=17), phrasal complexity (k=16), production integrity (k=11) and global complexity (k=13). Estimated (not observed) logarithm of coefficient of variation ratio (patients>controls) are given along with 95% credible intervals in parentheses. FTD=Formal Thought Disorder. nFTD: no-Formal Thought Disorder. FEP=First Episode Psychosis. SCZ=Established schizophrenia

Using Robust BMA, we found no or weak evidence for publication bias in all of the individual meta-analyses, with moderate to extreme evidence retained for domain-specific impairments in syntax (Table 2).

## 4. Discussion

To our knowledge, this is the first meta-analysis on the association between schizophrenia spectrum disorders and the use of grammar/syntax. BMA reveals extreme evidence in support of a global impairment across the domains of interest in schizophrenia, with the most robust effects being noted for comprehension of complex syntax and detection of errors followed by production length and integrity. This implies that people with schizophrenia spectrum disorders understand simpler sentences better, ignore syntactical errors, and speak in less sophisticated, shorter sentences that may not have a complete syntactic structure. The evidence favoring illness-related differences was moderately strong for global and phrasal complexity, even after taking between-studies heterogeneity and publication bias into account. Within the patient group, variability in grammar production/comprehension was higher than that of the healthy control group; this may occur in the presence of subgroups with varying degrees of impairment among patients. Taken together, a broad spectrum of grammatical impairment appears to be a key feature of schizophrenia.

Given the relatively modest sample sizes in individual studies (median patient n=32), our meta-analytic synthesis offers a more robust and representative effect size of grammatical impairment in SSD. Nevertheless, one limitation is our reliance on summary measures reported by authors instead of individual participant data. As 40% of case-control contrasts came from studies completed 20 years ago, we assessed (a priori) the likelihood of data availability to be low. Notable variation in study quality was noted, with representativeness across sexes and assessment languages being poor. Our synthesis is also limited by the diversity of variables used to define the domain-specific divergence; this likely accounts for the high heterogeneity observed in certain domains. Some overlap among the constructs of interest was noted (e.g., between global and phrasal complexity), while individual studies seldom reported the subject-level correlations among the various domains (especially between production and comprehension divergence), precluding our ability to test one of our pre-registered aims (but see the Supplement for the multivariate meta-analysis).

We also record notable variations in clinical sampling, with some studies focussing exclusively on those with FTD^102^. We found insufficient data to estimate the effect of FTD across all domains, and excluded studies that only compared FTD and non-FTD patient groups^125^. But our results indicate that grammatical impairment occurs irrespective of the presence of FTD. It is important to note that at an individual level, the degree of grammatical impairment is likely to be much higher among patients as it is influenced by comorbid developmental disorders and poor proficiency in a non-native language, both of which led to participant exclusion in the studies we identified. Furthermore, patients with more severe linguistic deficits often lack the capacity to provide written informed consent, making the effect size reported here a highly conservative estimate of the real-world complexities of grammatical impairment in schizophrenia. We make a set of recommendations for future studies in this regard (Supplemental Table 4).

One of the strengths of our review is the depth of our literature search - covering 50 years of work. In contrast to Ehlen and colleagues^4^ who recently “identified no studies evaluating syntax production in individuals with schizophrenia”, our search strategy located k=29 studies on syntax production. Furthermore, our Robust BMA analytical approach accounts for the uncertainty in heterogeneity and publication bias estimates and offers a comprehensive meta-analytic quantification of the overall magnitude of grammatical impairment in schizophrenia. The robust medium-to-large deficit in syntax production makes a strong case for including speech-based predictive analytics for early detection of schizophrenia, reinforcing prior^35,126^ and ongoing studies in this regard^127^.

Deficits in syntax, being a rule-based feature of language, are potentially remediable across the lifespan, and syntactic improvement may also affect other levels of linguistic processing (see Supplement Box 1). This has been shown in aphasic disorders with structured rehabilitation/education approaches (e.g., mapping therapy, syntax stimulation ^128,129^) or via targeted cognitive training (e.g., working memory^130^). By demonstrating evidence for a small-to-medium-sized increase in inter-individual variability in syntactic deficit (especially for phrasal complexity and syntactic comprehension), our synthesis encourages pre-trial selection of patients for communicative remediation. In particular, for syntactic comprehension, the combination of a large effect-size deficit, low between-studies heterogeneity, and the possibility of finding highly impaired subgroups indicate its suitability as an outcome measure for linguistic intervention trials.

The neural and social interactional basis of the observed syntactic deficits warrants attention in future studies. Emerging arguments against the presence of specific neural substrates for syntax/combinatorial processing in human language ^131–133^, indicate that the syntactic aberrations in schizophrenia may underwrite divergence at other levels of language processing, especially semantic cognition; this remains to be seen. Our observation of a generalized syntactic deficit across patient samples argues against focusing exclusively on those with clinically detectable FTD in mechanistic studies of linguistic divergence in psychosis (see ^64,134,135^ for a similar argument).

Our estimate of overall syntactic impairment (*d*=0·87) is smaller than the generalized cognitive impairment reported in schizophrenia (*d*=1·2^136^). Studies included in our meta-analysis either excluded participants with notably low IQ or matched IQ between groups; thus, we cannot attribute the observed syntactic divergence to a generalized cognitive impairment. Unlike the constrained neuropsychological tests used to assess cognitive deficits, syntactic divergence (especially in production) reported here has been observed on the basis of narratives/conversations that occur in more natural contexts. Thus, grammatical impairments, often carried by patients without much self-awareness, are likely to have intrusive effects on one’s everyday social functions.

In conclusion, our meta-analysis substantiates the long-suspected role of grammatical aberrations in schizophrenia. The question of whether these deficits occur independently of lexico-semantic abnormalities or are part of a broader linguistic impairment remains unresolved. Nonetheless, the findings underscore the need for targeted interventions to address these linguistic differences. More general implications include the importance of adjusting verbal exchanges in therapeutic settings for schizophrenia.

## Data Availability

All data produced in the present work are contained in the manuscript

## Funding

L. Palaniyappan’s research is supported by the Canada First Research Excellence Fund, awarded to the Healthy Brains, Healthy Lives initiative at McGill University (through New Investigator Supplement) and Monique H. Bourgeois Chair in Developmental Disorders. He receives a salary award from the Fonds de recherche du Quebec-Sante. This work is supported by the FRQS Partenariat Innovation-Québec-Janssen (PIQ-J) initiative (#338282); Canadian Institutes of Health Research (CIHR) - Strategy for Patient-Oriented Research Priority Announcement (SPOR; Grant number PJK192157) and Project Grant (Grant number PJT195903); Wellcome Trust Discretionary Grant (226168/Z/22/Z)

## Conflicts of interest

L.P. reports personal fees from Janssen Canada, Otsuka Canada, SPMM Course Limited, UK, Canadian Psychiatric Association; book royalties from Oxford University Press; investigator-initiated educational grants from Sunovion, Janssen Canada, Otsuka Canada outside the submitted work.

## Contributors

L.P. conceptualized the study; D.E. and L.P. designed, searched, and extracted the data; L.P., Y.C., and Q.L. undertook the meta-analysis. L.P. and D.E. interpreted the findings and drafted the manuscript; All authors revised it critically for important intellectual content.

## Data Availability

All data that support the findings are provided as supplementary materials. Any further data requests can be made to the corresponding author.

